# 12 Novel Clonal Groups of *Leptospira* Infecting Humans in Multiple Contrasting Epidemiological Contexts in Sri Lanka

**DOI:** 10.1101/2020.08.28.20177097

**Authors:** Dinesha Jayasundara, Indika Senavirathna, Janith Warnasekara, Chandika Gamage, Sisira Siribaddana, Senanayake Abeysinghe Mudiyanselage Kularatne, Michael Matthias, JF Mariet, Mathieu Picardeau, Suneth Agampodi, Joseph Vinetz

**Affiliations:** Leptospirosis Research Laboratory, Department of Community Medicine, Faculty of Medicine and Allied Sciences, Rajarata University of Sri Lanka; Department of Microbiology, Faculty of Medicine and Allied Sciences, Rajarata University of Sri Lanka; Department of Biochemistry, Faculty of Medicine and Allied Sciences, Rajarata University of Sri Lanka; Department of Microbiology, Faculty of Medicine, University of Peradeniya, Sri Lanka; Department of Medicine, Faculty of Medicine and Allied Sciences, Rajarata University of Sri Lanka; Department of Medicine, Faculty of Medicine, University of Peradeniya, Sri Lanka; Yale University school of Medicine, New Haven, Connecticut, USA; Institut Pasteur, Biology of Spirochetes unit, Paris, France

## Abstract

Leptospirosis is a ubiquitous disease and a major clinical challenge owing to the multitude of clinical presentations and manifestations that are possibly attributable to the diversity of *Leptospira*, the understanding of which is key to study the epidemiology of this emerging global disease threat. Sri Lanka is a hotspot for leptospirosis with high levels of endemic disease as well as annual epidemics. We carried out a prospective study of *Leptospira* diversity in Sri Lanka, covering the full range of climatic zones, geography, and clinical severity. Samples were collected for leptospiral culture from 1192 patients from 15 of 25 districts in Sri Lanka over two and half years period. Twenty five isolates belonging to four pathogenic *Leptospira* species were identified: *L. interrogans, L. borgpetersenii, L. weilii*, and *L. kirschneri*. At least six serogroups were identified among the isolates: Autumnalis (6), Pyrogenes (4), Icterohaemorrhagiae (2), Celledoni (1), Grippotyphosa (2) and Bataviae (1). Seven isolates did not agglutinate using available antisera panels, suggesting new serogroups. Isolates were sequenced by Illumina. These data add 25 new core genome sequence types and were clustered in 15 clonal groups, including 12 new clonal groups. *L. borgpetersenii* was found only in the dry zone and *L. weilii* only in the wet zone. Acute kidney injury and cardiovascular involvement were seen only with *L. interrogans* infections. Thrombocytopenia and liver impairment were seen in both *L. interrogans* and *L. borgpetersenii* infections. The inadequate sensitivity of culture isolation to identify infecting *Leptospira* species underscores the need for culture-independent typing methods for *Leptospira*.

**Author Summary:** There is a huge diversity in pathogenic *Leptospira* species worldwide, and our knowledge of the currently circulating species is deficient owing to limited isolation and identification of *Leptospira* species from endemic countries. This prospective study reveals the wide pathogen diversity that causes human leptospirosis in Sri Lanka, representing four species, more than six serogroups, and fifteen clonal groups. Further, the different geographic and climatic zone distributions and clinical manifestations observed underscores the need for prospective studies to expand the molecular epidemiological approaches to combat leptospirosis

## Introduction

Leptospirosis is caused by a group of pathogenic *Leptospira* species of the phylum Spirochetes and is considered one of the commonest zoonotic diseases worldwide [1][2]. *Leptospira* spp. have the ability to colonize proximal convoluted tubules of kidney tissue of various mammals (including rodents), birds and marsupials, and the hosts excrete the bacteria to the environment via urine [3][4]. Humans are incidental hosts who acquire the disease by direct contact with urine or tissues of reservoir animals or, more frequently by indirect contact with contaminated water sources [5][6]. The number of cases due to leptospirosis is estimated to be 1.03 million annually worldwide, with 58,900 deaths [1]. The majority of tropical countries in Oceania, southeast Asia, the Caribbean region, central and eastern sub-Saharan Africa, and south Asia are estimated to have substantial morbidity and mortality that is attributable to leptospirosis.[7]

Understanding the diversity of infecting *Leptospira* has been a major global focus, especially in recent years. The phenomenal changes in *Leptospira* classification backed by next-generation sequencing methods and whole-genome sequencing have led to the identification of 43 new *Leptospira* species during the period 2018 to 2020 [8–11]. In addition, the more robust classification of *Leptospira* strains beyond the species level using core-genome multi-locus sequence typing (cgMLST) [9] and single nucleotide polymorphism typing methods has rapidly expanded our knowledge of the molecular epidemiology of *Leptospira*. However, the goal of reducing the global burden of this deadly disease will require enhanced understanding of pathogen types and applications and linkage to disease distribution, transmission, clinical presentations, and outcomes.

The global leptospirosis disease burden study [1] has highlighted Sri Lanka as a hyperendemic country with an estimated morbidity of 300.6 and mortality of 17.98 per 100,000 population per year. The disease incidence tends to be higher during the rainy seasons, i.e., the southwest and northeast monsoons. Cases, however, are not confined exclusively to the wet zone and are reported in the dry zone as well, where the majority of residents are engaged in farming activities. Outbreaks have also occurred in the dry zone following extreme weather events like flooding [12].

As in many other endemic countries, understanding *Leptospira* diversity in Sri Lanka is limited because of a lack of knowledge of the circulating pathogenic species and serovars. Studies that utilized culture-based isolation of *Leptospira* species were carried out in Sri Lanka during the period from 1950 to 1970 in the wet zone only. Several pathogenic strains of the species *L. interrogans* [13][14][15], *L. borgpetersenii* [16], *L. kirschneri* [17], and *L. santarosai* were detected during that time [18][19]. Since the 1970s, no culture based isolation studies were reported until 2018, when two human isolates belonging to *L. interrogans* were recovered from the wet zone [20]. Despite the availability of next-generation sequencing methods for many years, whole-genome sequencing data for Sri Lankan isolates were not available until recently [21].

A systematic review published in 2016 revealed the large diversity of *Leptospira* strains in Sri Lanka based on historical data [19]. Being an island with a high leptospirosis disease burden makes Sri Lanka an ideal location to study pathogen diversity linked with epidemiological and clinical patterns of the disease. Low-passage isolates from human sources with high-resolution genetic typing in a place with high pathogen diversity would enhance our global knowledge of leptospirosis. This study was designed to provide a comprehensive understanding of the circulating pathogenic *Leptospira* species and serotypes responsible for human leptospirosis in Sri Lanka, covering different clinical presentations and geographical locations as well as epidemic and endemic disease over a period of two and half years.

## Methods

The present study was embedded in a larger clinical-epidemiological study on leptospirosis, in Sri Lanka and the study protocol was published elsewhere [22]. Specific details related to *Leptospira* diversity and methods in brief are given here.

### Study setting

This study was carried out from June 2016 through January 2019 at several locations in Sri Lanka that differed with respect to mean temperature, rainfall, elevation, ecology, human activities, and leptospirosis endemicity. The main data collection sites were the Teaching Hospital Anuradhapura (THA) and Teaching Hospital Peradeniya (THP). THA is in the dry zone located at low elevation with low humidity, high temperature, large rice paddy fields, water reservoir–based irrigation systems, and low endemicity for the disease. THP is in the wet zone located at high elevation, low temperature, rainfall-based farming activities, and high endemicity. Samples for *Leptospira* diversity assessment were collected through two approaches. First, as a part of the main study described in Agampodi et al. 2019, prospective data and sample collection was done in THA and THP. In addition, during an outbreak of leptospirosis in 2017, we set up the same procedure at Base Hospital Avissawella and Provincial General Hospital Rathnapura from June to September. These two wet-zone areas have high endemicity, representing low and intermediate elevations. As a part of the service component of this study, we offered diagnostic services to all requesting physicians and also collected additional culture samples. This resulted in sample collection from District General Hospital Kegalle, Base Hospital Karawanella, Sri Jayawardanapura General Hospital, and General Hospital Polonnaruwa (GHP) again representing different geographical locations. These study sites (Fig 1) represent seven districts belonging to four provinces of the country, and the patients who visited these hospitals came from all nine provinces.

**Fig 1.**
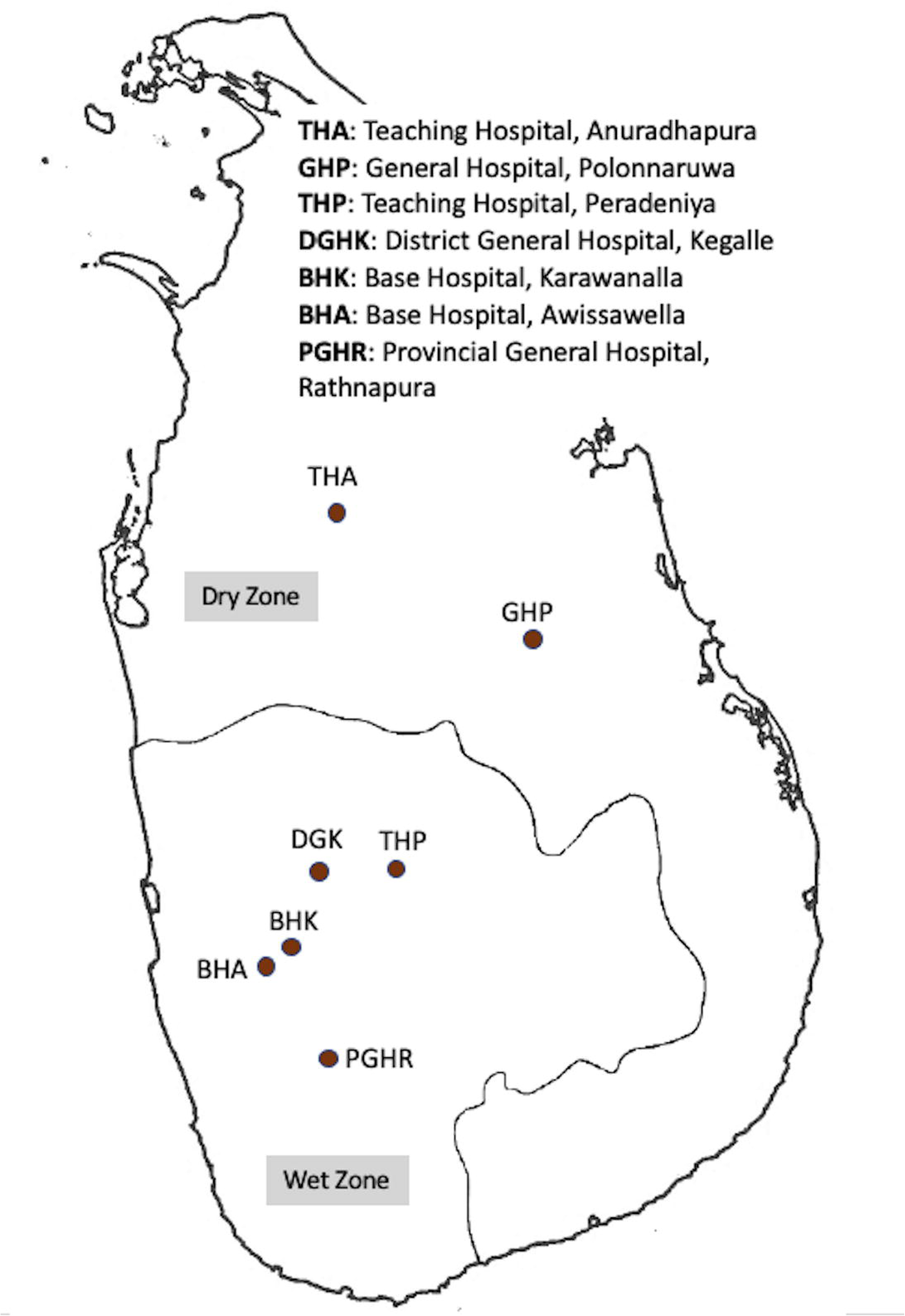
Locations of the seven hospitals involved in the study

### Study samples

Culture collection was done from three types of patients. Acute undifferentiated febrile (temperature >38°C) patients who presented to adult wards (age >13 years) of THA, THP, Provincial General Hospital Rathnapura (PGHR) and Base Hospital Avissawella (BHA) (both outpatient department and hospitalized patients) were included as possible cases of leptospirosis. A possible case was defined as any acute undifferentiated febrile patient with headache, mayalgia and prostration. Probable cases of clinical leptospirosis which were defined as those who were having the classical clinical features of leptospirosis with an exposure history, were included from GHP. Culture samples from Sri Jayawardanapura General Hospital and Base Hospital Karawanella were included only if they came from clinically confirmed cases of leptospirosis. These cases were defined according to the surveillance case definition for leptospirosis set by epidemiology unit of Sri Lanka[23]. Physician-diagnosed probable or definite acute bacterial meningitis or lower respiratory tract infections (e.g., consolidated lobar pneumonia), traumatic or post-operative fever per physician discretion, fever owing to nosocomial infections, and any patient with confirmed diagnosis as a cause for the fever were excluded. Epidemiological data were collected from each patient using a fully structured, interviewer administered questionnaire which was described in detail in the study protocol paper published elsewhere[22]

### Sample collection and isolation of Leptospira

Blood (7 ml) was collected into EDTA tubes from all eligible patients. Bedside inoculation of 2 and 4 drops (100–400 µl) was done into two tubes with 9 ml Ellinghausen-McCullough-Johnson-Harris (EMJH) semisolid medium with added antibiotics (5-fluorouracil and neomycin). These cultures were kept at room temperature (usually 28– 32°C) until transfer to the Leptospirosis Research Laboratory of the Faculty of Medicine and Allied Sciences, Rajarata University, Sri Lanka, and then incubated at 30°C until the cultures become positive or for 6 months. Samples from THA were transferred on the same day to the research laboratory whereas other samples collected from distant places were transferred within 2 days of collection.

In brief, EMJH semisolid media were prepared by adding 2.3 g of EMJH base (Difco), 1.5 g bacteriological agar, and 100 mg sodium pyruvate into 785 ml distilled water and adjusting the pH to 7.4. The media were autoclaved, and once cooled to ∼50°C, 100 ml *Leptospira* enrichment media and 100 ml fetal bovine serum were added. To suppress the growth of possible contaminating bacteria, 5-fluorouracil (100 µg/ml, final) and neomycin (25 µg/ml) were added. Each inoculated medium was inspected by taking approximately 50µl of volume into a clean glass slide after mixing the culture tubes well by inverting several times. Prepared slides were examined under 40X objective of a dark-field microscope to check for the presence of motile spirochetes; this was done initially after 3 weeks and then on a monthly basis. However, samples were inspected before 3 weeks if quantitative PCR of the corresponding whole-blood sample indicated a positive reaction.

The procedure for qPCR on clinical samples is described elsewhere in the published protocol paper[22]. Culture tubes were inspected for consecutive 4 months before reporting as negative. When positive growth was detected, subcultures were made into liquid and semisolid media, and an aliquot was fixed with 5% dimethyl sulfoxide and stored at –80°C. Isolates were subcultured in liquid media once in 2 weeks and on semisolid media once in 3 months. Certain isolates required weekly subculture into liquid media to maintain viability. None of the isolates became contaminated during the subculture process, although two positive original clinical samples were contaminated with bacilli. For those two samples, subcultures were made into liquid media and subsequently filtered through a 0.2 µm pore-size microfilter to overcome the problem of contamination.

### Next-generation sequencing, cgMLST and phylogenetic tree

DNA was extracted from culture using the PureLink Genomic DNA Mini kit (Invitrogen, Dublin, Ireland) and Wizard Genomic DNA Purification Kit (Promega, Southampton, UK) according to manufacturer instructions. NGS was performed using Nextera XT DNA Library Preparation kit and the NextSeq 500 sequencing systems (Illumina, San Diego, CA, USA) at the Mutualized Platform for Microbiology (P2M) at Institut Pasteur. The data were analyzed using CLC Genomics Workbench 9 software (Qiagen, Hilden, Germany). cgMLST typing was performed for strain taxonomy using a scheme based on 545 highly conserved genes with BIGSdb (http://bigsdb.pasteur.fr/leptospira), and a phylogenetic tree was generated using cgMLST with Interactive Tree of Life v3, and GrapeTree [24] was used to visualize the core genomic relationships among the isolates and the previously reported Sri Lankan isolates [9][25][26]. Clonal Groups (CG) is defined as a group of cgMLST allelic profiles differing by no more than 40 allelic mismatches, out of 545 gene loci, from at least one other member of the group.

### Serotyping of new isolates

Serotyping of newly isolated *Leptospira* strains was done at the Pasteur Institute, France. Microscopic agglutination test using a standard battery of rabbit antisera raised against 24 reference serovars representing the main serogroups was used for this study [27][28].

### Ethics statement

Written informed consent was obtained from all patients prior to sample collection. This study is approved by the Ethics Review Committee of the Faculty of Medicine and Allied Sciences, Rajarata University of Sri Lanka. Protocol No. ERC/2015/18

## Results

From June 2016 through January 2019, we acquired blood cultures from 1192 patients. Patients were from 14 districts of Sri Lanka, representing all 9 provinces (Fig 2).

**Fig 2.**
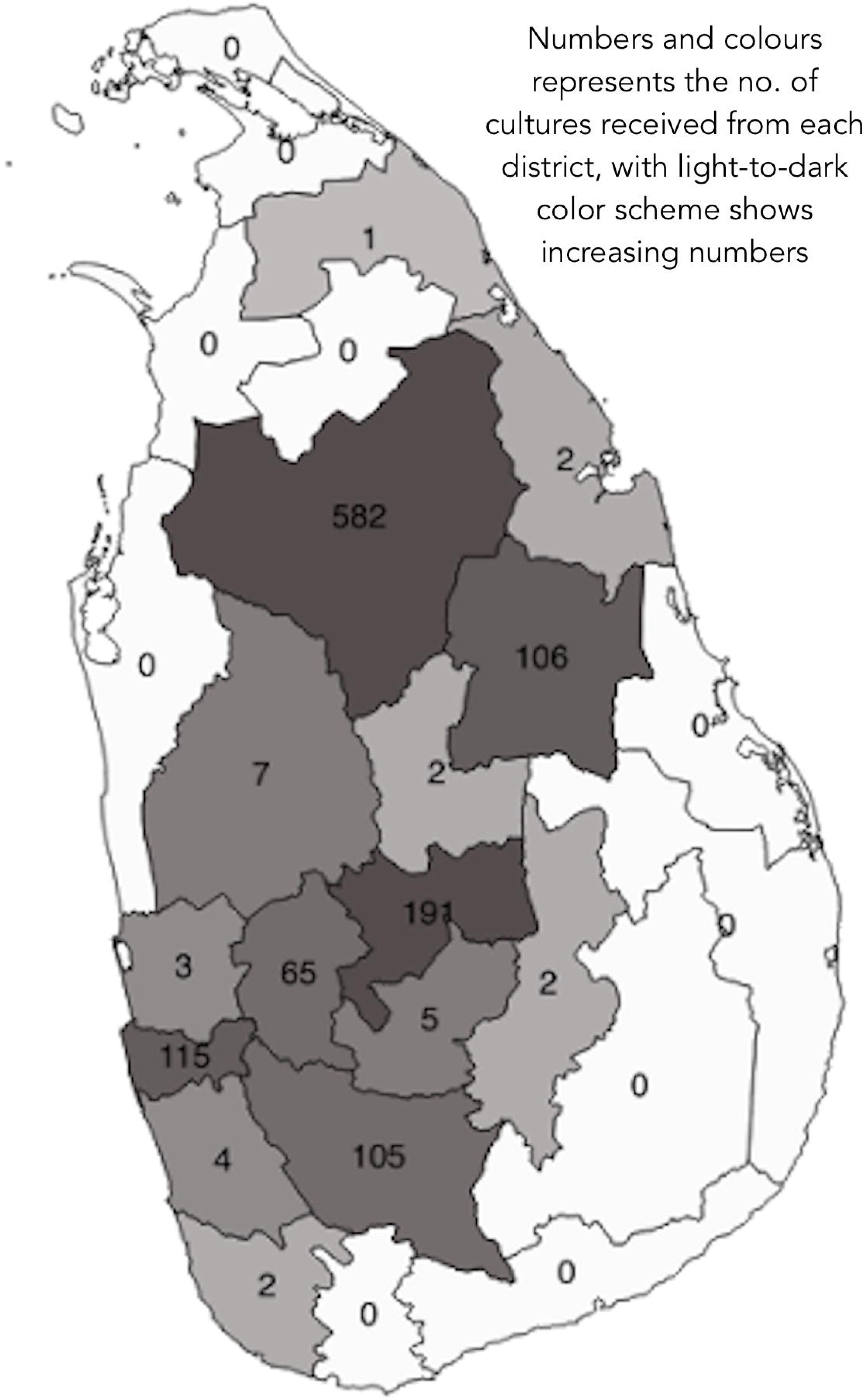
Distribution of probable exposure sites/residence of the patients recruited for the study.

The majority of patients were male (n = 985, 82.6%). The mean age of the sample was 43.4 years (SD 14.7). Most of the cultures (96%) were obtained from hospital inpatients, and the remaining 4% (48 cultures) were from outpatients. Of the 1192 cultures, 80 (6.7%) were received from hospitals where only typical clinical cases of leptospirosis were sampled (from Sri Jayawardanapura General Hospital, Base Hospital Karawanella and Provincial General Hospital Rathnapura). Another 107 (9.0%) were from GHP, where patients represented only probable cases of leptospirosis. The remaining 1005 patients (84.3%) were from THA, THP, Base Hospital Awissawella and Provincial General Hospital Rathnapura, representing patients who presented with acute undifferentiated fever.

Of the 1192 patients, 25 isolates had been identified by January 2019. Among the acute undifferentiated febrile patients, 1.5% (16/1047) had culture-positive leptospirosis; among the probable and clinically confirmed cases of leptospirosis, culture positivity was 4.7% (5/107) and 5.0% (4/80), respectively. The incubation period required to detect positive growth (assessed with dark-field microscopy) varied between 1 and 17 weeks. For each patient, both 2 drops and 4 drops of blood-inoculated media gave positive results. The median incubation period was 15 weeks for the first 9 isolates and was 6 weeks for the remaining 16 isolates (Sup.figure1).

*Leptospira* was isolated from one female and 24 male patients who presented with fever. Only three patients were from outpatient departments, and the rest were inpatients. Of the three outpatients, two were later admitted to a hospital owing to increased disease severity.

cgMLST analysis revealed that the 25 isolates represented four species: *L. interrogans* (15 isolates, 60%), *L. borgpetersenii* (7 isolates, 28%), *L. weilii* (2 isolates, 8%), and *L. kirschneri* (1 isolate, 4%) The isolates which were classified using core genome MLST genotyping scheme based on clusters created at the 40-mismatch level revealed the clonal group identity of them. A clonal group was defined using a single linkage clustering with a threshold set at 40 allelic mismatches. The predominant clonal group (CG) was CG267 (All 7 *L. borgpetersenii* isolates among the 25 total isolates). This was followed by CG266 (3/25), CG10 (2/25), and CG263 (2/25) of *L*.*interrogans*. The two *L. weilii* isolates clustered in different clonal groups (CG262, CG264). Interpretable data for serogroup assay was available for only 19 isolates. Seven samples resulted in no agglutination, probably owing to new, previously unreported serogroups/serovars. The assay revealed that the 19 seropositive isolates represented at least six serogroups, namely Autumnalis, Pyrogenes, Icterohaemorrhagiae, Grippotyphosa, Celledoni, and Bataviae. Table 1 shows the species identity, serogroup status and distribution of clonal groups among the 25 isolates

**Table 1.** Putative species, serogroups, cgMLST and clonal groups for the *Leptospira* isolates

| Strain ID | Species | Serogroup | cgMLST | CG |
| --- | --- | --- | --- | --- |
| FMAS_KW1 | *L. interrogans* | Pyrogenes | 784 | 10 |
| FMAS_KW2 | *L. interrogans* | Autumnalis | 631 | 291 |
| FMAS_AW1 | *L. interrogans* | Autumnalis | 555 | 74 |
| FMAS_RT1 | *L. weilii* | No agglutination | 556 | 262 |
| FMAS_AW2 | *L. interrogans* | Autumnalis | 567 | 269 |
| FMAS_AW3 | *L. interrogans* | Pyrogenes | 557 | 9 |
| FMAS_RT2 | *L. interrogans* | Autumnalis | 569 | 271 |
| FMAS_PD1 | *L. interrogans* | Pyrogenes | 558 | 263 |
| FMAS_PD2 | *L. weilii* | Celledoni | 559 | 264 |
| FMAS_KG1 | *L. interrogans* | Bataviae | 560 | 265 |
| FMAS_KG2 | *L. interrogans* | Pyrogenes | 561 | 263 |
| FMAS_AP1 | *L. interrogans* | Autumnalis | 562 | 266 |
| FMAS_AP2 | *L. borgpetersenii* | No agglutination | 563 | 267 |
| FMAS_AP3 | *L. borgpetersenii* | No agglutination | 575 | 267 |
| FMAS_AP4 | *L. borgpetersenii* | No agglutination | 564 | 267 |
| FMAS_AP5 | *L. interrogans* | Pyrogenes | 565 | 10 |
| FMAS_AP6 | *L. interrogans* | Pyrogenes | 785 | 321 |
| FMAS_AP7 | *L. interrogans* | Autumnalis | 786 | 266 |
| FMAS_PN1 | *L. borgpetersenii* | No agglutination | 787 | 267 |
| FMAS_PN2 | *L. interrogans* | Icterohaemorrhagiae | 788 | 322 |
| FMAS_PN3 | *L. interrogans* | Autumnalis | 789 | 266 |
| FMAS_PN4 | *L. borgpetersenii* | No agglutination | 790 | 267 |
| FMAS_AP8 | *L. borgpetersenii* | No agglutination | 791 | 267 |
| FMAS_AP9 | *L. borgpetersenii* | No agglutination | 792 | 267 |
| FMAS_PN5 | *L. kirschneri* | Grippotyphosa | 793 | 323 |

**cgMLST**- core genome Multi Locus Sequence Typing, **CG**: clonal group defined as a group of cgMLST allelic profiles differing by no more than 40 allelic mismatches, out of 545 gene loci, from at least one other member of the group

A phylogenetic tree was constructed from the cgMLST data for the 25 isolates together with available data for previous reported nine (09) local isolates and currently circulating pathogenic species worldwide. Fig 4 shows the phylogenetic tree which was constructed with the cgMLST data.

**Fig 3.**
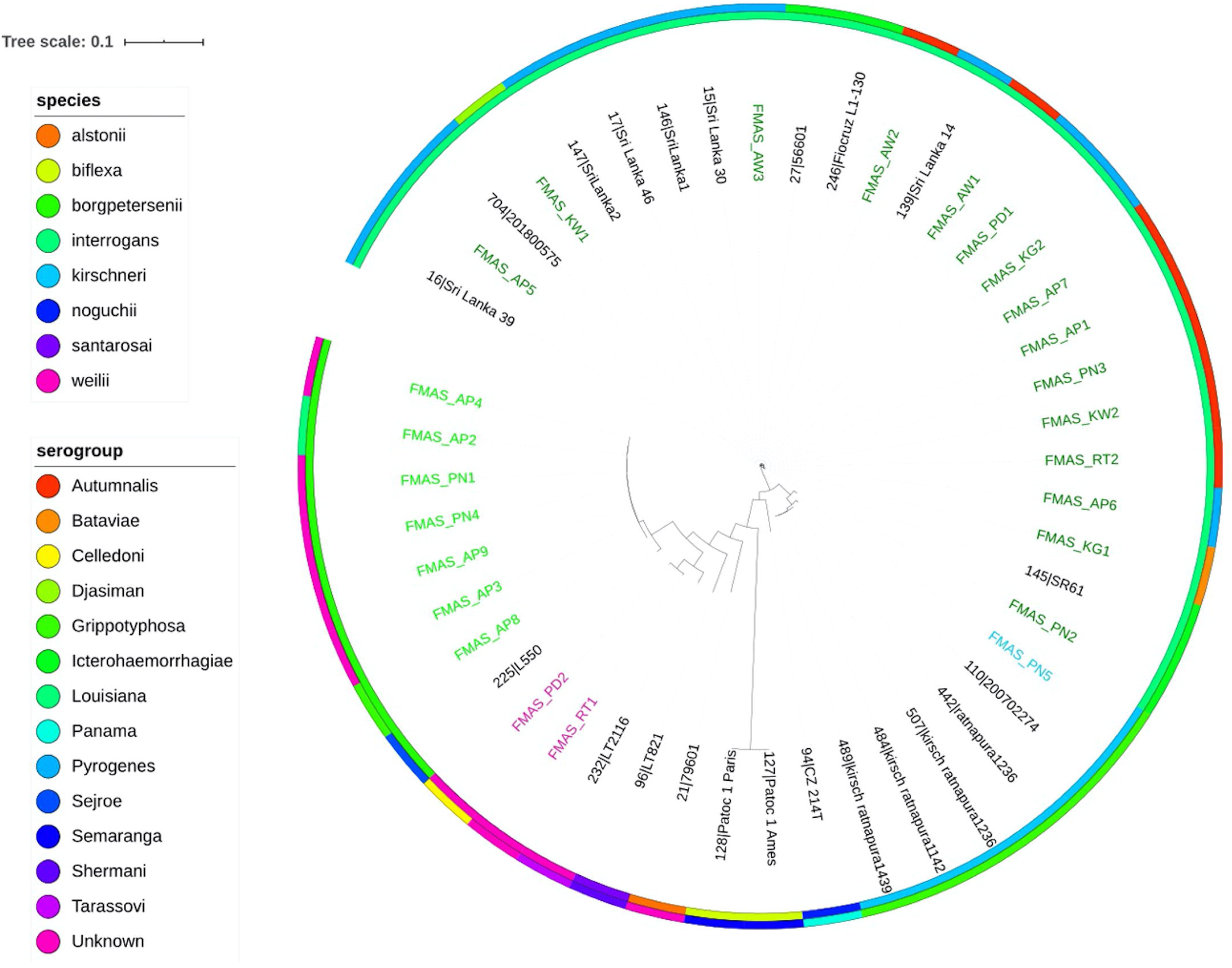
Phylogenetic tree showing the distribution of species and serogroups of the isolates from the present study along with the previously reported Sri Lankan isolates and others species.

**Fig 4.**
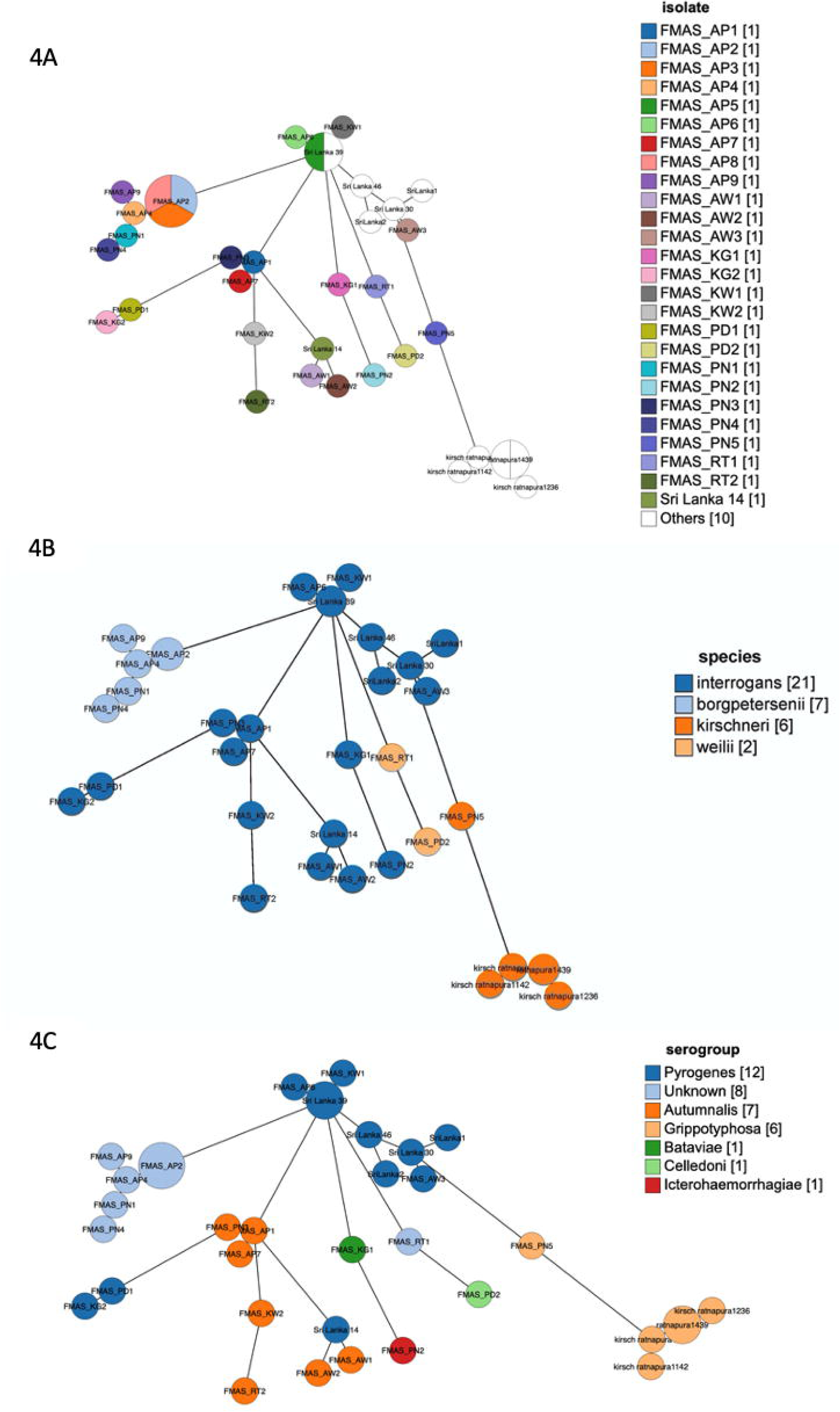
Genome GrapeTree showing the core-genome relationship among the 25 new and 9 previously isolated *Leptospira* strains from Sri Lanka

Most of the clonal groups found in the local isolates were unique and not found in other countries. FMAS_AP2, FMAS_AP3 and FMAS_AP8 of *L*.*borgpetersenii* shows clear core genomic relatedness forming a single cluster from the other four isolates of the same species (Fig 4A). These three isolates were from patients in the same district of dry zone. The previously reported four isolates of *L. kirschneri* from Rathnapura was distinct from newly isolated *L*.*kirschneri* from Polonaruwa, even though they are in the same arm. Two clear clusters of interograns could be observed. In one cluster AP7,PN3 AP1 of serogroup Autamnalis from dry zone shows significant core genome relatedness and clustered together. The other cluster was formed with FMAS_AP6, FMAS_KW1, FMAS_AP5 and isolate Sri Lanka 39 of Pyrogenes serogroup. These isolates represent both dry and wet zones. There’s genomic relatedness between FMAS_AP5 of dry zone and Sri Lanka 39 which is an old isolate recovered from wet zone. Another close association was observed between FMAS_KG2 and FMAS_PD1 which are from wet zone and of serogroup Pyrogenes. Two L. welli isolated from Rathnapura(RT1) and Peradeniya (PD2) have not shown a significant core genomic relation. (metadata for the GrapeTree is included in Sup.Table 1)

Culture-positive leptospirosis patients were distributed widely in the study areas (Fig 5). *L. borgpetersenii* was exclusively isolated from patients in areas of the dry zone at low elevation, with hot and dry conditions. The single isolate of *L. kirschneri* was from the same setting. In contrast, *L. weilii* was isolated from patients in the wet zone, whereas *L. interrogans* was isolated from patients in all geographical areas (Fig 5). The serogroup distribution also revealed a specific pattern for all nonagglutination isolates, which mainly were from the dry zone at low elevation. The most frequent serogroup Autumnalis, was observed in all geographical settings. All *L. borgpetersenii* isolates were in CG267, the majority of which failed to agglutinate with rabbit sera (some slightly agglutinated with serovars Grippotyphosa and Louisiana).

**Fig 5.**
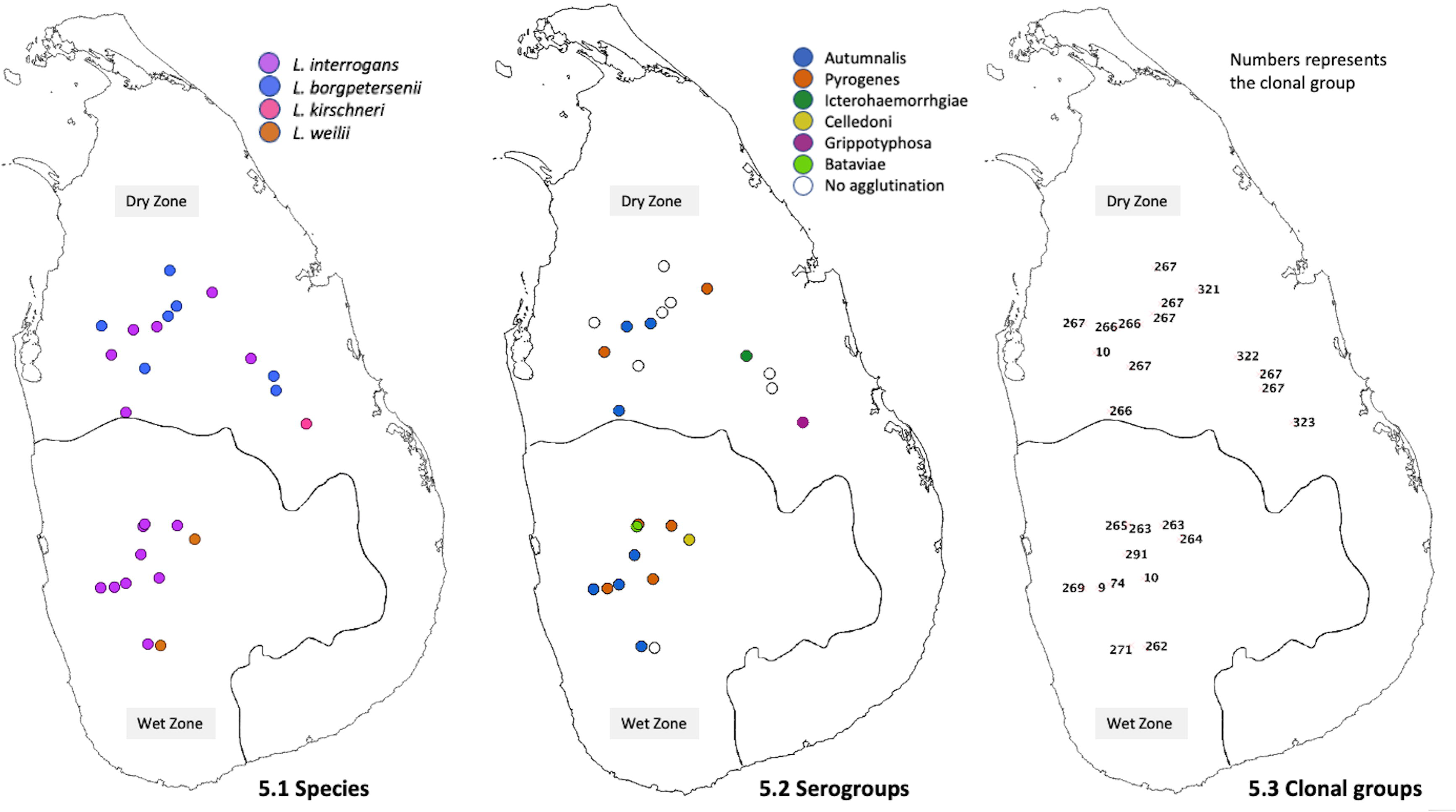
Geographic distribution of *Leptospira* species, serogroups and clonal groups

### Clinical profile of culture-positive patients

Complete demographic and clinical profiles for the culture-positive patients are included as a supplementary data (Sup. Table 1). All culture-positive patients were later contacted and/or visited to collect additional data. The additional data were collected mainly to identify exact type of exposure if possible and residing places of patients preceding the illness. In addition, diagnosis cards of these patients were also traced to extract any missing data during hospital stay. Each patient’s clinical records were also retrieved from the corresponding hospital. No fatalities were reported for patients with culture-positive leptospirosis. Among hospitalized patients, the median duration of hospital stay was 4 days (interquartile range, 3–5 days), and the longest stay was 12 days followed by 9 days (both patients had acute kidney injury and required hemodialysis). Another two had renal involvement with elevated serum creatinine but not acute kidney failure. Twelve patients had elevated serum glutamic-oxaloacetic transaminase and serum glutamic-pyruvic transaminase, and three other patients had elevated serum bilirubin. Thrombocytopenia was common in those 12 patents, with a platelet count <100,000 per microliter. Two patients who underwent hemodialysis also had cardiac involvement with hypotension. Each patient who had a severe complication was infected with *L. interrogans*. Infection with *L. interrogans* or *L. borgpetersenii* was associated with thrombocytopenia and liver involvement.

## Discussion

This is the first study to report information on the diversity of pathogenic *Leptospira* species in Sri Lanka and probably one of the few prospective studies on *Leptospira* disease diversity in literature representing all geographical regions of an entire country. In addition, we report here the first isolation of *L. weilii* in Sri Lanka, the existence of which was suggested based on molecular studies of clinical samples [29]. This study also provides the first evidence of serogroups Celledoni and Bataviae circulating in Sri Lanka. Moreover, with the exception of *L. santarosai*, our study identified all pathogenic *Leptospira* species that were reported to have existed in Sri Lanka during the 1960s and 70s (as reviewed by Naotunna et al. in 2015), confirming the breadth of *Leptospira* diversity in Sri Lanka [19]. Recent reports indicated that *L. interrogans, L. borgpetersenii* and *L. kirschneri* are also the most common circulating species in other tropical regions of the world [30][31][32][33][34][35]. However, the geographical distribution of certain *Leptospira* species is limited, such as *L. santarosai*, which has been mainly reported in South America [36][37][38][39], and *L. weilii* in Asia [35][40][41][42]. Only limited reports have described the existence of *L. weilii* outside Asia, where cattle and rodents are the dominant reservoir hosts [42][43][44][45].

We identified 15 distinct clonal groups of *Leptospira*, underscoring the diversity of pathogenic *Leptospira* circulating in Sri Lanka. The unique identity of most clonal groups among the local isolates emphasizes the significance of conserved reservoir hosts and serovars in an island. Genomic relatedness wasn’t observed between most of old Sri Lankan isolates with these new isolates probably due to changes in the bacterial genome or due to emergence or introduction of new strains. The distribution of certain clonal groups revealed a geographical demarcation between dry and wet zones. A single clonal group status and restriction to dry zone of all *L borgpetersenii* isolates together with core genomic relatedness among three isolates from same district are interesting observations. These might reflect similarities in climate, reservoir hosts and environmental conditions in a particular geographical area. Clustering of *L*.*interrogans* into several clonal groups and shared genomic relatedness across different geographical areas could possibly be due to diversity of reservoir hosts and adoptability of the species to different environmental conditions. These observations might possibly suggests that different environmental drivers of leptospirosis operate in distinct ways for different pathogenic species and their serovars in these climatic zones. The predominance of *L. borgpetersenii* and *L. kirschneri* over *L. interrogans* has been reported in both humans and cattle in the African continent and nearby islands such as Mayotte, with possible cattle-to-human transmission [46][47][48]. Moreover, certain rat species also excrete serovars of *L. borgpetersenii* [49][50]. Notably, areas where *L. borgpetersenii* was found in our study were all in the dry zone where cattle and buffalo are commonly used in paddy farming activities.

Majority of the total study population and culture positive patients were males and this is likely to be due to exposure to possible risk factors as males engage in outdoor activities frequently than women. This pattern of involvement is seen across most of the published studies worldwide[1][7][51][52]. According to available clinical data renal, hepatic, haematological and cardiovascular complications are observed in infections with *L*.*interrogans*. This is consistent with most published data worldwide[53][54][55]. In contrast either haematological complications alone or with hepatic involvement was observed in infection with the single clonal group of *L*.*borgpetersenii*. However pulmonary complications were not observed among the 25 culture positive patients.

In our present study, *L. interrogans* was the predominant species identified in patients residing in the wet zone. Emergence of a single dominant clone that caused an outbreak of leptospirosis following a flood was reported in Thailand] [32]. In contrast with that study, we observed diverse clonal groups of *L. interrogans* as the cause of human leptospirosis during floods, probably owing to disease transmission from several different reservoir hosts.

The diversity of circulating serogroups in Sri Lanka that has been known since the 1960s has been preserved, and 19 isolates belong to 6 different serogroups, and the non-agglutinated cultures might reflect other unidentified serovars other serovars. Serological and molecular assays done on veterinary field has already identified the role of rodents, cattle and dogs as reservoir hosts in Sri Lanka[56][57][58]. However the observed diversity in serogroups offers evidence for a wider range of reservoir hosts despite the fact that Sri Lanka is a small island.

Although culture isolation is required for in-depth molecular epidemiological studies, our present study highlights the constraints faced for culture isolation of *Leptospira*, for which both a high level of skill and procedural optimization are required. *Leptospira* spp. are fastidious organisms, and their growth requirements differ from those of many other bacterial genera. *Leptospira* tend to have a relatively long incubation period, as the lag period during *in vitro* culture range from days to several weeks [59]. Other culture-isolation studies have reported an incubation period of ∼3 weeks, but with a wide range of duration [46][60][61]. We attribute the relatively lengthy incubation period of 15 weeks required for the first nine cultures in our study partly to lack of to detect if any scanty growth of *Leptospira* during the first phase of the study. However it’s also possible that those isolates were fastidious and required lengthy incubation periods. Isolation is essential for genomic and vaccine studies pertaining to *Leptospira*, and for that purpose a specific skill set is mandatory. Although culture isolation has 100% positive predictive value for diagnosing leptospirosis, its sensitivity has been consistently <10% in most studies [32][62][63]. Similarly, our study yielded low sensitivity, which can be attributed to a few possible causes: Because the study population consisted of patients with acute febrile illness, their fever may have been caused by another unrelated illness; Use of antibiotics prior to culture; also, performing blood cultures during the late phase of illness and infection with fastidious *Leptospira* spp. may account for the low sensitivity during culture isolation.

This large collection of pathogenic *Leptospira* isolates from clinical samples will be a great addition to the global knowledgebase for leptospirosis. Whole-genome sequencing and genomic analysis of this set of isolates will reveal the pathogenic diversity and evolution of *Leptospira* species, in comparison to archived *Leptospira* isolated from Sri Lanka more than 50 years ago. The first three isolates from this study are already published[21] and available in NCBI genome database (https://www.ncbi.nlm.nih.gov/genome/?term=Leptospira+interrogans).

Whole genome sequencing and comparative genomic analysis of this collection will facilitate ongoing studies on identifying the putative virulent genes, pathogenic mechanisms with specific host adaptations, horizontal gene transfer mechanisms, and microbial resistance as shown in studies on the diversity and epidemiology of other microorganisms [64][65][66][67][68][69].

## Data Availability

All genomic data are available from NCBI (accession numbers are submitted). De-identified patient data are in the supplementary table.

## Acknowledgement

This work was supported by The National Institute of Allergy and Infectious Diseases of the National Institutes of Health, Award Number U19AI115658. The content is solely the responsibility of the authors and does not necessarily represent the official views of the National Institutes of Health. The funding body had no role in the design of the study or the collection, analysis, or interpretation of data or in writing the manuscript and publication. We would like to thank Ms. Thilakanjali Gamage, Mr. K.M.R. Premathilaka, Mr. S.K. Senevirathna, and Mr. Milinda Perera for technical assistance, Mr. Shalka Srimantha and Ms. Chamila Kappagoda for culture maintenance and laboratory support, and Dr. Muditha Abeykoon, Dr. Chamida Wickramasinghe, and Dr. Shanika Gamage for additional culture collections. We also thank all the physicians and healthcare staff in the various participating hospitals, team of core facility P2M (Institut Pasteur, Mutualized Platform for Microbiology) for genomic sequencing and members of the National Reference Center for Leptospirosis (Institut Pasteur) for technical assistance with the cultures of *Leptospira*.

---

**Fig 1. Locations of hospitals used in this study and the ecological zones of Sri Lanka**

**Fig 2. Distribution of febrile patients by probable exposure sites/ residence.**

**Fig 3. Phylogenetic Tree showing the species and serogroup distribution of the isolates from the present study (labelled as FMAS_) with previously reported Sri Lankan isolates and others species.**

**4A: isolates, 4B: species, 4C:serogroups. New isolates are having the prefix FMAS**

**Fig 4. Genome GrapeTree showing the core-genome relationship among 25 new and 11 previously isolated Leptospira strains from Sri Lanka.**

**Fig 5. Geographical distribution of *Leptospira* species, serogroups and clonal groups**

**Sup.Fig.1: Distribution of incubation period for culture isolation among 25 *Leptospira* cultures**

**Sup.Table 1: Clinical and ddemographic profile of culture positive patients**

